# Neuroanatomical correlates of genetic risk for obesity in children

**DOI:** 10.1101/2022.06.07.22275937

**Authors:** Filip Morys, Eric Yu, Mari Shishikura, Casey Paquola, Uku Vainik, Gideon Nave, Philipp Koellinger, Ziv Gan-Or, Alain Dagher

## Abstract

Obesity has a strong genetic component, with up to 20% of variance in body mass index (BMI) being accounted for by common polygenic variation. Most genetic polymorphisms associated with BMI are related to genes expressed in the central nervous system. At the same time, higher BMI is associated with neurocognitive changes. However, the direct link between genetics of obesity and neurobehavioral mechanisms related to weight gain is missing. Here, we use a large sample of participants (n>4,000) from the Adolescent Brain Cognitive Development cohort and investigate how genetic risk for obesity, expressed as polygenic risk score for BMI (BMI-PRS), is related to brain and behavioral differences in adolescents. In a series of analyses, we show that BMI-PRS is related to lower cortical volume and thickness in the frontal and temporal areas, relative to age-expected values. Relatedly, using structural equation modeling, we find that lower overall cortical volume is associated with higher impulsivity, which in turn is related to an increase in BMI 1 year later. In sum, our study shows that obesity might partially stem from genetic risk as expressed in brain changes in the frontal and temporal brain areas, and changes in impulsivity.

## 1 Introduction

Obesity is consistently associated with alterations in brain anatomy [1–10]. These changes can be detected by magnetic resonance imaging (MRI) and affect both grey and white matter. Numerous mechanisms have been proposed to explain these findings.

Brain morphometry may underpin behavioral traits that predispose to obesity [11– 14]. Indeed, obesity is heritable and polygenic [15–18] - twin studies show that the heritability of body mass index (BMI) is 70%, and large-scale meta-analyses of genome-wide association studies (GWAS) have identified over 700 risk alleles [15, 18–20]. Polygenic risk scores for BMI (BMI-PRS), representing the sum of genetic risk per individual, account for 5-15% of variance in BMI proper [18, 21]. It has been suggested that the risk alleles act in the brain to cause a behavioral phenotype that renders individuals prone to positive calorie balance and weight gain [17, 22, 23]. This phenotype may involve satiety and hunger signaling in the hypothalamus, but may also include the trait uncontrolled eating, which has been linked to function of brain systems implicated in learning and memory, stress, motivation, and executive control [14, 15, 23, 24].

Conversely, it is also known that obesity can change the brain. Chronically, adiposity is associated with a metabolic syndrome that includes hypertension, hyperlipidemia, and insulin resistance, which can cause brain atrophy over time [25]. Even over shorter time-spans, positive calorie balance may lead to adaptive brain changes. Studies of diet-induced obesity in rodents show widespread alterations in synaptic density and neuronal composition, occurring in parallel with or even prior to weight gain [26–33]

Disentangling the chain of causality between brain anatomy and obesity is difficult. The approach used here is to estimate the effect of polygenic risk for obesity on brain anatomy in young children. This would be expected to limit the effect of potential confounds due to metabolic syndrome and have a better chance to identify the inherited neural endophenotype that renders individuals vulnerable to weight gain. We focus on genetic potential for adult obesity, as the goal of childhood obesity prevention should be avoidance of adulthood obesity as it is causally linked with negative health outcomes [34]. Here, we analyze how genetic risk for adulthood obesity is associated with brain structure, executive function, impulsivity, and 1-year BMI change in a sample of 4157 children aged 9-11 years.

## 2 Materials and methods

### 2.1 Participants – main sample (ABCD)

We used data from the Adolescent Brain Cognitive Development Cohort (ABCD), a longitudinal, multi-site study from the USA [35–37]. We excluded participants with outlier BMI values (below 10 kg/m^2^ or above 50 kg/m^2^) [11]. We only included participants with full neuroimaging data and participants who passed all quality control checks. The final sample consisted of 4,157 children of European descent (mean age=10 years, SD=0.5 year; mean BMI=17.94 kg/m^2^, SD=3.30 kg/m^2^; mean weight=35.60 kg, SD=8.58 kg). At the present time PRS calculation is only possible in individuals of European descent as GWAS were conducted in this population. BMI values were converted to standard deviation scores (BMI SDS), which are z-scores derived from age and sex of each participant, based on Centers for Disease Control and Prevention growth charts [38] using the ‘childsds’ package in R (mean BMI SDS=0.20 kg/m^2^, SD=1.09 kg/m^2^). 1-year BMI SDS change values were calculated as the difference between BMI SDS at a follow-up appointment and BMI SDS at the time of brain and behavioral data collection, hence higher values represent a BMI SDS increase.

### 2.2 Neuroimaging data – main sample (ABCD)

Data were collected using 3T magnetic resonance imaging (MRI) scanners of different manufacturers (Siemens, General Electric, and Philips) at 22 different sites. Data collection was harmonized across all acquisition sites by using standardized hardware (e.g., head coils) and adjusting acquisition sequences for each scanner manufacturer. All imaging protocols can be found elsewhere [35]. We used cortical thickness, cortical volume, and fractional anisotropy data provided by the ABCD initiative [39]. Cortical thickness and volume data for each parcel of the Desikan-Killiany (DK; 68 parcels) atlas [40] were obtained using FreeSurfer 5.3.0 [41] after correcting for gradient nonlinearity distortions. Custom scripts were used to obtain fractional anisotropy data for 35 major white matter tracts segmented using AtlasTrack [42]. Visual inspection of processed data was conducted to ensure that only images with no processing errors were included in the dataset. We used quality check values provided by the ABCD Study (pass/fail) to only include participants who passed quality control in our final sample. Prior to all but brain age analyses, we used ComBat software to remove site variability from cortical thickness, volume, and FA data [43]. In addition, we scaled all volumetric measures by total intracranial volume.

### 2.3 Polygenic risk score calculation

We assessed genetic risk for obesity using body mass index polygenic risk scores (BMI-PRS). BMI-PRS was calculated using PRSice-2 [44] with pruning and thresholding of BMI GWAS summary statistics from an independent data sample (https://www.ebi.ac.uk/gwas/downloads/summary-statistics)[18]. First, variants in the GWAS summary statistics were pruned based on linkage disequilibrium of variants within 250kb and r^2^>0.1. Then, the p-value threshold where the PRS was most correlated with phenotypical BMI in the investigated dataset (ABCD) was chosen for PRS calculation. We normalized all BMI-PRS for downstream analyses. GWAS sample and ABCD sample are independent.

### 2.4 Participants – brain age sample (PING)

To place the anatomical findings in the context of expected neurodevelopmental stage, we used a separate sample of similar age. We created a predicted brain age model to which we compared each ABCD participant’s grey matter measures. To this end, we used the Pediatric Imaging, Neurocognition, and Genetics (PING) data [45]. This dataset encompasses 1493 children aged 3-20 years collected across multiple sites in the US. For our analysis, we selected a subsample with brain measures and age data available (n=781). The average age of this sample was 13 years (SD=5 years), and range was 3 to 18 years.

### 2.5 Neuroimaging data – brain age sample (PING)

Data were collected using 3T MRI scanners from GE, Siemens, or Philips. Data collection was harmonized across 10 study sites by adjusting acquisition sequences. Details of data acquisition and processing can be found in [45]. Here, we used grey matter volume and thickness measures provided by the PING consortium, as derived from FreeSurfer [41].

### 2.6 Executive function and impulsivity measures

To relate our findings to behavioral measures, we used executive function and impulsivity indices, as both were previously related to obesity [46, 47]. Impulsivity was assessed using the child version of the Urgency, Premeditation, Perseverance, Sensation Seeking, Positive Urgency scale (UPPS-P [48, 49]). Here, we selected positive and negative urgency measures as they were previously associated with eating behavior [50, 51]. To assess executive function, similarly to [52], we calculated a composite score based on the results of five tests, namely the Flanker inhibitory control and attention test, the dimensional change card sort test, the picture sequence memory test, the list sorting working memory test, and the pattern comparison processing speed test [52–56](correlations with the composite score: Flanker test: 0.64; card sort test: 0.71; picture memory test: 0.58; working memory test: 0.59; processing speed test: 0.72; all p-values<0.001). Age corrected scores were used and a composite score for each participant was derived by averaging the five standardized scores.

### 2.7 Data analysis

Statistical analyses were conducted using R (v. 3.6.1; [57]).

#### 2.7.1 Relationship between BMI-PRS and phenotypical BMI SDS

We first conducted a proof-of-concept analysis to investigate whether BMI-PRS was related to measured BMI SDS. To this end, we used regression analysis with BMI SDS as an outcome variable and BMI-PRS, first 20 genetic principal components (to control for population stratification), age, sex, interaction of age and sex, and study site as predictor variables.

#### 2.7.2 Relationship between brain volume and BMI-PRS

Next, we explored the relationship between genetic risk for obesity (BMI-PRS) and brain measures of interest – cortical thickness, cortical volume, and fractional anisotropy. We ran separate permutation-based regression analyses (10,000 permutations) for each cortical parcel and white matter tract. We used BMI-PRS as a predictor of brain measures, while accounting for age, sex, parental education, parental income, parental marital status, child’s education, and first 20 genetic principal components. We used Benjamini-Hochberg correction to adjust for multiple comparisons [58]. This was applied separately to cortical thickness, cortical volume, and white matter measures. Brain plots were prepared using ‘fsbrain’ package in R [59].

#### 2.7.3 Cortical architectonic types

To gain a better understanding of the cortical areas affected by BMI-PRS, we investigated how they relate to brain cortical profiles of the organization of cortical types along sensory processing hierarchies (i.e. idiotypic, unimodal, heteromodal, or paralimbic cortex). We used cortical architectonic maps representing the sensory-fugal gradient [60, 61] and parcellated them using the Desikan-Killiany atlas [40]. Next, we calculated total cortical grey matter volume and average cortical thickness of each of the cortical types for each subject and regressed these measures against BMI-PRS using previously described covariates (permutation-based regression, 10,000 permutations). We then calculated effect sizes (partial eta squared) to compare which architectonic types were most strongly associated with BMI-PRS.

#### 2.7.4 Brain age analysis

Brain maturation in adolescence is associated with changes in grey and white matter. We therefore related the neuroanatomical effects of BMI-PRS to brain age in the ABCD sample based on a model from a different dataset. This model was based on the PING sample and used grey matter measures while correcting for intracranial volume. We then used the model to estimate the brain age of participants in the ABCD sample. The model was derived using linear regression with 10-fold cross validation. Due to low sample size in the PING dataset, the model was based on global measures of brain structure: cortical grey matter volume, and average cortical thickness, rather than individual DKT parcels. Prior to the analysis, sex and study site effects were removed from the measures from both ABCD and PING using linear regression. Model performance was assessed using root mean square error (RMSE). We initially included linear and quadratic terms in the model, however, since the model did not improve significantly with quadratic terms, we decided to only use linear terms in the final estimation.

Brain age for each participant in the ABCD dataset was estimated based on imaging features and the model described above. For each participant, we calculated a difference between chronological age and brain age – delta age [62, 63]. Finally, we correlated delta age with BMI-PRS (corrected for first 20 genetic principal components). Because delta age is correlated with chronological age, we used Spearman’s partial correlation analysis to regress out the effects of chronological age [62].

#### 2.7.5 Structural equation model

To pool our results in one model and investigate how BMI-PRS can affect BMI SDS change via brain and behavioral changes, we used a structural equation model (SEM) with the following measures of interest: BMI-PRS, global brain measures – cortical thickness, cortical volume, and FA – executive function and impulsivity, and 1-year change in BMI SDS. Data were first residualized to remove variance related to age, sex, parental education, parental marital status, household income, child’s education, study site, and first 20 genetic principal components. Additionally, BMI SDS change was residualized for baseline BMI SDS to make sure that the effects found in the SEM were not due to correlations of our variables of interest with baseline BMI SDS.

Using the lavaan package in R (version 0.6-5 [64]) we created an SEM, where we hypothesized that BMI-PRS would affect average cortical thickness, total grey matter volume, and average FA. These measures were then hypothesized to affect impulsivity and executive function, which would in turn affect BMI SDS. We allowed for residual correlations between impulsivity and executive function measures, and between brain measures.

We estimated the model using maximum likelihood estimation with pairwise missing values exclusions and robust standard errors. Model fit was assessed using root mean square error of approximation (RMSEA), standardized root mean square residual (SRMR), and comparative fit index (CFI).

## 3 Results

### 3.1 Relationship between BMI-PRS and phenotypical BMI SDS

BMI-PRS was significantly positively associated with BMI SDS (standardized beta coefficient estimate=0.263, t=17.610, F(45,4111)=10.39, 95% confidence intervals: 0.255-0.319, p<0.001; Figure 1a). BMI-PRS accounted for 7.2% variance in BMI SDS beyond the first 20 genetic principal components, age, sex, study site, and age*sex interaction. As for the adults in Locke et al. [15], the BMI-PRS SNPs were normally distributed in our population, and the effect of risk SNPs on BMI was additive (Figure 1a). BMI SDS, BMI-PRS, and genetic principal components strongly differed by study site (Figure 1b-d), hence all the results presented here are corrected for study site and genetic principal components unless otherwise stated.

**Figure 1.**
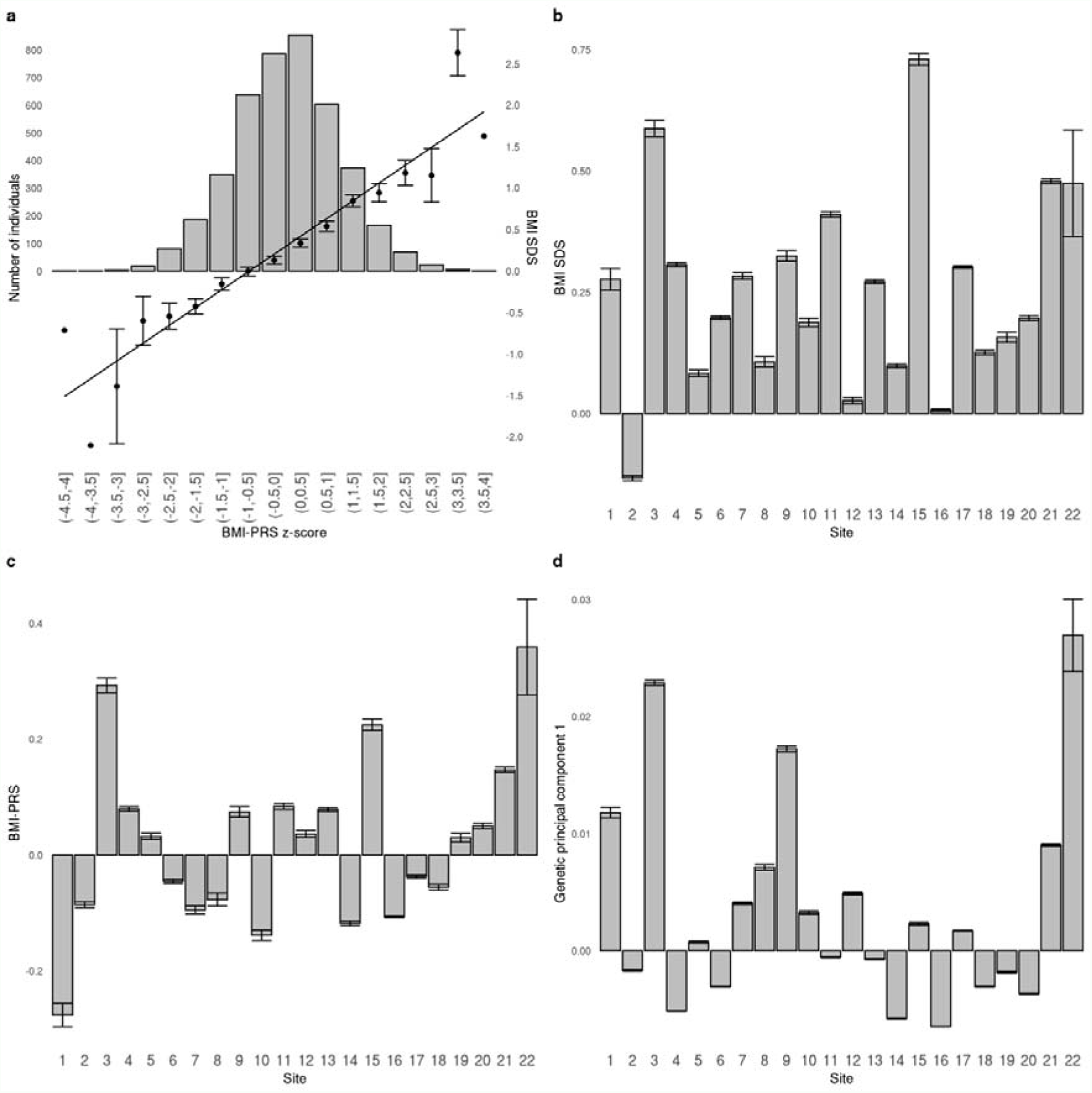
**a** Relationship between BMI-PRS z-score and BMI SDS (right axis) overlayed on a histogram (left axis). The histogram represents the number of participants in each BMI-PRS z-score bin; **b** relationship between BMI SDS and study site; **c** relationship between BMI-PRS and study site; **d** relationship between the first genetic principal component and study site. BMI-PRS: body mass index polygenic risk score. BMI SDS: body mass index standard deviation score.

### 3.2 Brain-PRS relationship

BMI-PRS was negatively associated with cortical volume in several brain areas, predominantly in the frontal and temporal lobes (Table 1, Figure 2a). The strongest associations were visible in the left precentral and right superior frontal gyri. BMI-PRS was also associated with lower cortical thickness in the frontal and temporal areas (Table 1, Figure 2b). We did not find any significant associations between BMI-PRS and white matter fractional anisotropy.

**Table 1.**
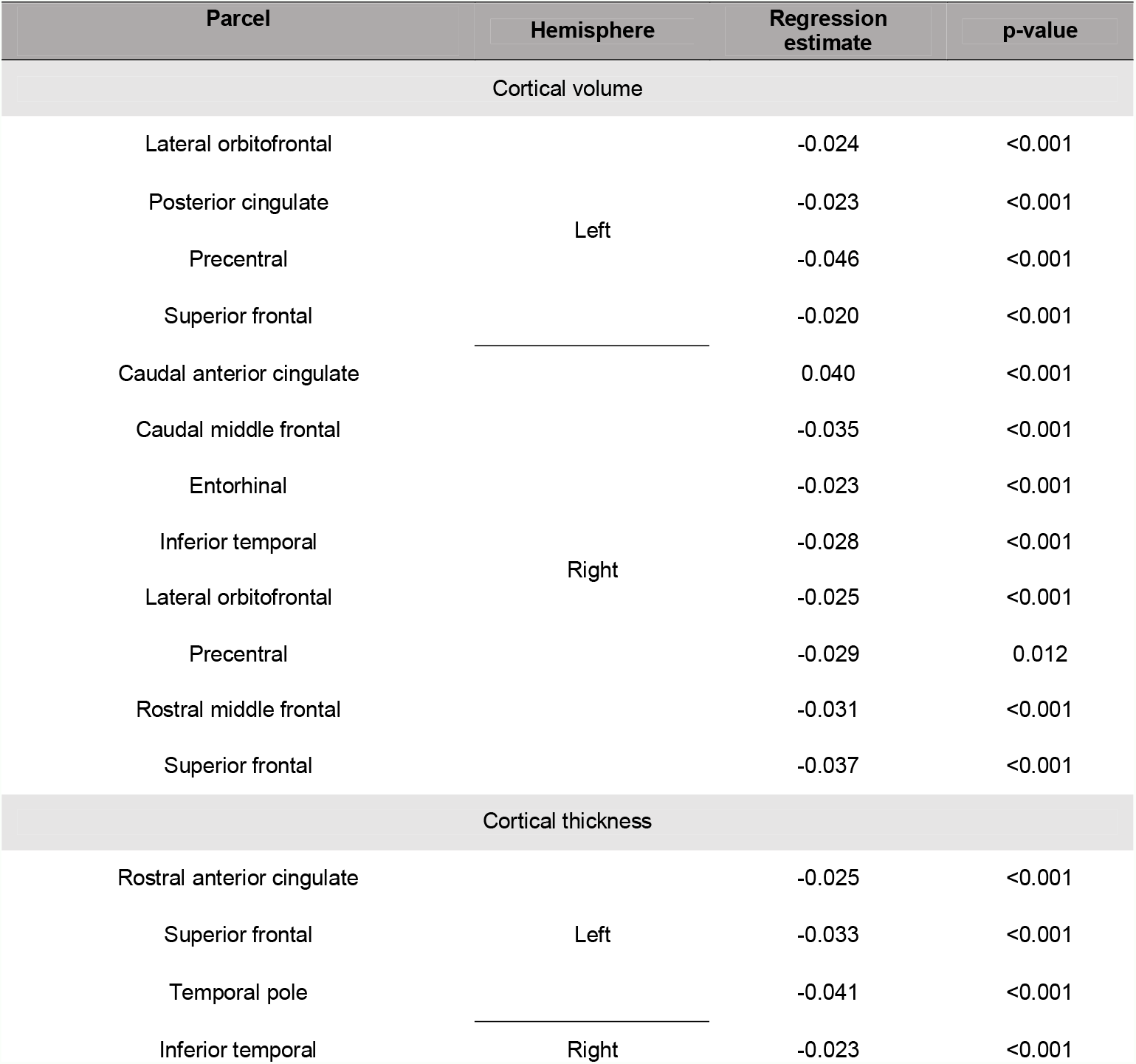
Relationship between BMI-PRS, cortical thickness and cortical volume.

**Figure 2.**
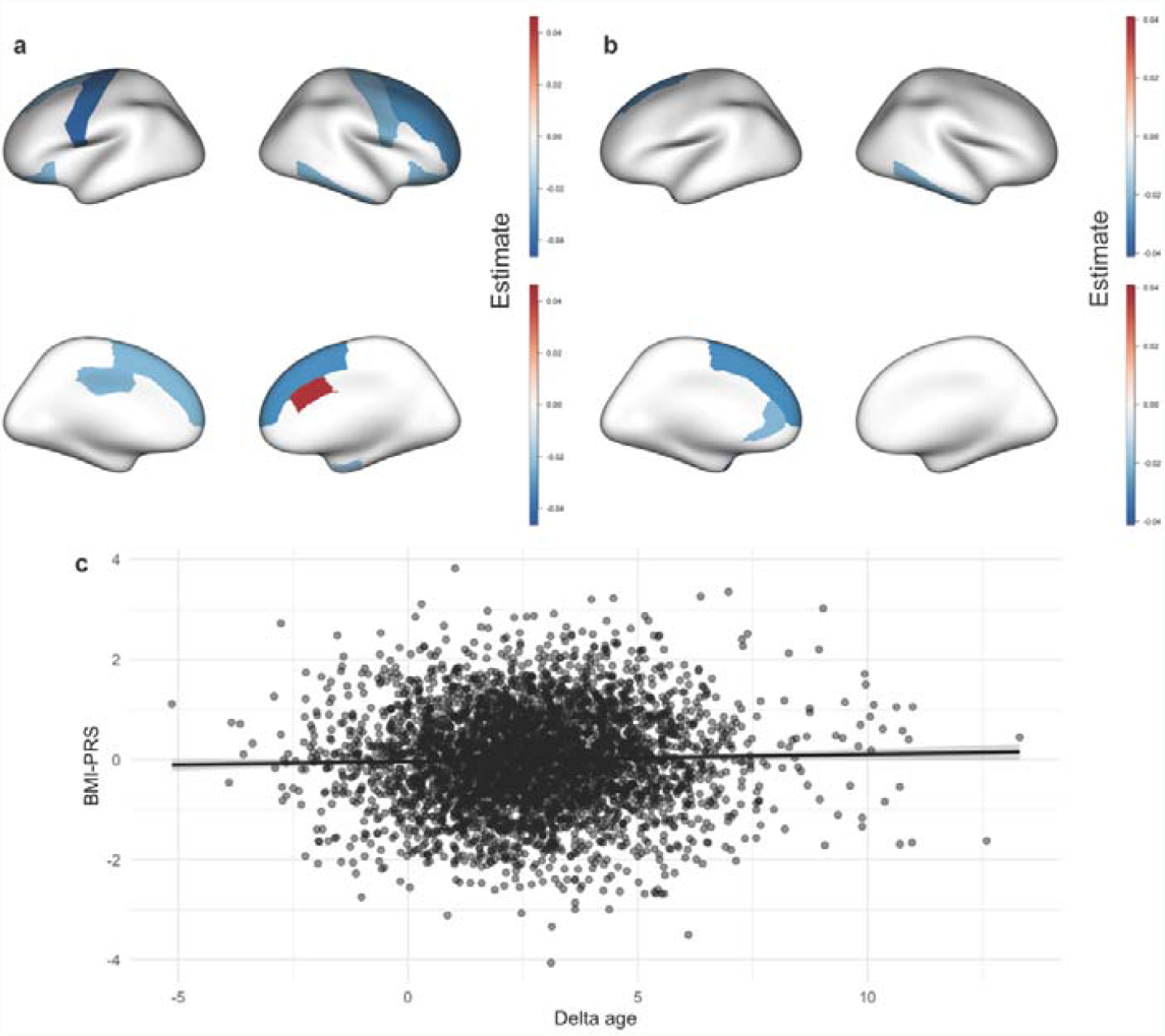
**a** Relationship between BMI-PRS and cortical volume; **b** relationship between BMI-PRS and cortical thickness; **c** relationship between BMI-PRS and predicted brain age.

### 3.3 Cortical architectonic types

To investigate whether cortical volume and thickness changes related to BMI-PRS belong to specific cortical architectonic types defined by Mesulam [60], we calculated total cortical volume and average cortical thickness for each of the types (idiotypic, unimodal, heteromodal, and paralimbic) per subject and investigated their association with BMI-PRS. Total cortical volume of the heteromodal and unimodal cortex, but also average cortical thickness of the heteromodal cortex were significantly negatively associated with BMI-PRS (Table 2).

**Table 2.** Association between cortical thickness/volume for each cortical architectonic type and BMI-PRS.

| Cortical type | Partial $\eta^2$ | Regression estimate | p-value |
| --- | --- | --- | --- |
| Cortical volume |  |  |  |
| **Idiotypic** | 0.0001 | -0.103 | 1.000 |
| **Unimodal** | 0.0008 | -0.203 | <0.001 |
| **Heteromodal** | 0.0012 | -0.252 | <0.001 |
| **Paralimbic** | 0.0002 | -0.104 | 0.106 |
| Cortical thickness |  |  |  |
| **Idiotypic** | 0.0002 | 0.009 | 0.959 |
| **Unimodal** | 0.0001 | -0.008 | 0.574 |
| **Heteromodal** | 0.0004 | -0.012 | 0.033 |
| **Paralimbic** | 0.0002 | -0.009 | 1.000 |

### 3.4 Brain age analysis

The grey matter-based brain age model returned an RMSE of 3.49 years. Higher age was related to lower cortical thickness and volume. Partial correlation analysis showed that grey matter-related delta age in the ABCD dataset was significantly positively related to BMI-PRS (r=0.032, p=0.042; Figure 2c). This indicates that higher genetic risk for obesity is associated with cortical thinning and volume decreases, relative to age-expected values.

### 3.5 Structural equation model

The structural equation model returned a good fit (RMSEA=0.041, CFI=0.979, SRMR=0.020; Χ^2^=55.349, p<0.001). We found significant negative associations between BMI-PRS and global cortical volume, between global cortical volume and both negative and positive urgency, a significant positive association between fractional anisotropy and executive function, and a significant positive association between negative urgency and 1-year BMI SDS change (Table 3, Figure 3). Together, these associations illustrate a potential pathway by which BMI-PRS could affect BMI SDS change via brain alterations that are related to higher impulsivity.

**Table 3.** Structural equation model results. BMI-PRS body mass index polygenic risk score. BMI SDS body mass index standard deviation score.

| Outcome variable | Predictor | Estimate | Standard error | z-value | p-value |
| --- | --- | --- | --- | --- | --- |
| Cortical volume | BMI-PRS | -0.027 | 0.012 | -2.242 | 0.025 |
| Cortical thickness | BMI-PRS | -0.011 | 0.013 | -0.818 | 0.414 |
| Fractional anisotropy | BMI-PRS | 0.003 | 0.011 | 0.229 | 0.819 |
| Executive function | Cortical volume | -0.028 | 0.023 | -1.251 | 0.211 |
|  | Cortical thickness | -0.027 | 0.019 | -1.424 | 0.154 |
|  | Fractional anisotropy | 0.100 | 0.020 | 5.041 | 0.000 |
| Positive urgency | Cortical volume | -0.156 | 0.068 | -2.282 | 0.022 |
|  | Cortical thickness | 0.051 | 0.059 | 0.860 | 0.390 |
|  | Fractional anisotropy | 0.030 | 0.060 | 0.503 | 0.615 |
| Negative urgency | Cortical volume | -0.139 | 0.062 | -2.249 | 0.025 |
|  | Cortical thickness | 0.007 | 0.054 | 0.128 | 0.898 |
|  | Fractional anisotropy | -0.087 | 0.057 | -1.537 | 0.124 |
| BMI SDS increase | Executive function | -0.007 | 0.010 | -0.735 | 0.462 |
|  | Positive urgency | 0.002 | 0.004 | 0.472 | 0.637 |
|  | Negative urgency | 0.010 | 0.004 | 2.440 | 0.015 |

**Figure 3.**
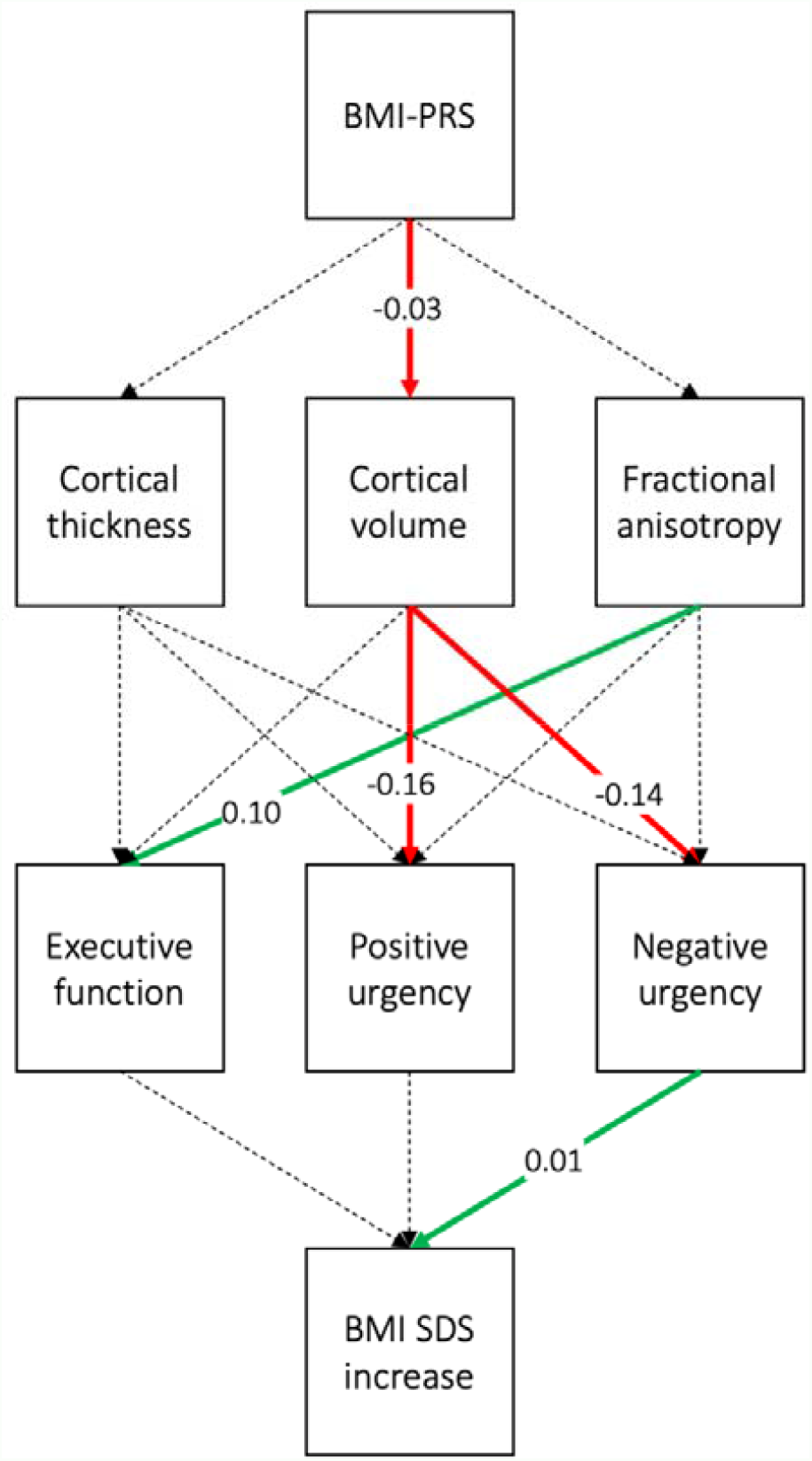
Schematic representation of the structural equation model. Significant associations are marked in red/green, non-significant associations are marked with dashed lines. BMI-PRS body mass index polygenic risk score. BMI SDS body mass index standard deviation score.

## 4 Discussion

Obesity is partially heritable and highly polygenic, and most of the genetic associations identified act in the central nervous system [15–17, 19]. Environmental factors are thought to interact with a vulnerable brain to favour weight gain, a theory supported by evidence that genetic predisposition to obesity is exacerbated in an obesogenic environment [17, 65]. Studies in adults have linked elevated BMI to executive function, impulsivity, and neuroanatomical changes, all of which have been argued to impact feeding behaviour [5, 14, 25, 46, 50–52, 66]. However, the direction of causality is unclear as chronic adiposity also leads to widespread changes in grey and white matter, and consequent cognitive changes [67]. We aimed to reduce the role of this possible confound by studying the effect of genetic risk rather than BMI *per se* in young children.

The risk alleles for obesity were normally distributed in our population, and the number of risk alleles per individual was linearly proportional to BMI SDS. Both were also observed in the Locke et al. GWAS study of 330,000 adults [15]. This suggests that the genetic disposition for obesity is already influencing BMI before age 10. We further found effects of BMI-PRS on grey matter morphometry, supporting the proposition that these genes exert their obesogenic effects via the brain.

High BMI-PRS was related to differences in grey matter volume and thickness relative to age-expected values. More specifically, BMI-PRS was associated with reduced grey matter volume and cortical thickness in bilateral fronto-temporal areas. Some of these regions are associated with executive function and have long been implicated in the control of food intake and impulsivity in adults [24, 68–70]. Our findings raise the possibility that cognitive control and impulsivity are also implicated in childhood body weight. Indeed, changes in brain morphometry were also associated with both a composite score for executive function and personality measures of impulsivity (positive and negative urgency), while impulsivity was related to an increase in BMI SDS. The fact that the heteromodal and unimodal levels of sensory processing hierarchies were most affected by BMI-PRS is consistent with impulsive individuals exhibiting a non-normative maturation of the heteromodal regions [60, 71]. It is also in line with our previous findings relating those heteromodal and unimodal regions to BMI in adults [72]. Overall, this suggests that impulsivity and fronto-temporal cortical morphometry are linked to obesity in young children, and that the effect may mediate some of the genetic risk for obesity. Importantly, grey matter change has been associated with obesity itself in adults, not obesity risk, although there it is thought to reflect damage secondary to metabolic consequences of adiposity [25]. While we cannot rule out such an effect in 9–10-year-olds studied here, a more likely explanation may be that polygenic risk for obesity is associated with brain development.

The influence of impulsivity on body weight and lack of influence of executive function on body weight in this population warrants some discussion. The links between obesity and reduced executive function have been established repeatedly in adults and children, even in the ABCD sample [46, 52]. Indeed, uncontrolled eating, a heritable trait associated with BMI that encompasses many aspects of food intake regulation [14], is correlated to lower performance on executive function tests in adults and higher impulsivity [24, 73], although these effects are not always consistent [14, 74]. Most authors suggest that cognitive control plays a role in food intake via decision making about portion size, food choice, and the like, however it is unlikely that this type of cognitive control is relevant in children. As for impulsivity, the links with BMI are also well-established, even in children [50, 75–78]. Studies in infants and children have shown that genetic risk for obesity manifests predominantly as increased hunger, reduced satiety, food responsiveness, or uncontrolled eating [23, 79–82], some of which could be related to impulsivity [83]. This would be in line with our findings that show genetic influence on BMI SDS change via brain and impulsivity measures.

A major limitation of our study and, perhaps, all studies investigating genetic risk for obesity in the ABCD sample, is the fact the BMI and BMI-PRS were not equally distributed over different study sites. BMI and BMI-PRS were also correlated with genetic principal components that reflect population stratification and that also differed by study site. While it is likely that the site differences in BMI-PRS also reflect population stratification, the differences in BMI might be related to other phenomena, such as different obesogenic environments, which is not considered as a confounding factor in our analysis. Thus, removing variance associated with genetic principal components could also remove some of the true associations between BMI-PRS or BMI and neurobehavioural data. Therefore, our study might not reflect all true associations between the investigated obesity-related features. Further, our study is a predominantly a cross-sectional one and so does not allow to properly investigate causal associations between BMI-PRS, brain structure, executive function, and impulsivity. Finally, we also did not directly investigate any aspect of eating behavior.

The current research could support genetic risk for obesity as causally related to developmental brain changes, leading to excess weight accumulation via higher impulsivity, or eating-related traits that are heritable and already expressed in infancy. This research contributes to a better understanding of neural mechanisms of obesity in adolescence, and could inspire future strategies for obesity prevention. Because BMI-PRS can be calculated early in life, individuals at a high risk for obesity could be identified and targeted by interventions that are, e.g., aimed at decreasing impulsivity, which could lead to beneficial health outcomes in the future.

## Data Availability

Data used in this manuscript are available from the Adolescent Brain Cognitive Development study representatives

https://abcdstudy.org/

## 5 Acknowledgments

This work was supported by a Foundation Scheme award to AD from the Canadian Institutes of Health Research, by computing resources from Calcul Quebec (www.calculquebec.ca) and Compute Canada (www.computecanada.ca), the Funai Foundation for Information Technology fellowship to MS, and by a Fonds de recherche du Québec - Santé (FRQS) foreign post-doctoral training award to FM. UV has been funded by Estonian Research Council’s personal research funding start-up grant PSG759. This work was also financially supported by Parkinson Canada, the Canadian Consortium on Neurodegeneration in Aging (CCNA), and the Canada First Research Excellence Fund (CFREF), awarded to McGill University for the Healthy Brains for Healthy Lives (HBHL) program. ZGO is supported by the Fonds de recherche du Québec - Santé (FRQS) Chercheurs-boursiers award, and is a William Dawson Scholar.

## 6 Conflict of interest

Disclosure Statement: FM, EY, CP, MS, UV, and AD have nothing to declare. ZGO has received consulting fees from Idorsia, Ono Therapeutics, Neuron23, Handl Therapeutics, UBC, and Bial Biotech Inc. None of these companies were involved in any parts of preparing, drafting, and publishing this study.

## Notes

### Competing Interest Statement

FM, EY, CP, MS, UV, GN, PK, and AD have nothing to declare.
ZGO has received consulting fees from Idorsia, Ono Therapeutics, Neuron23, Handl Therapeutics, UBC, and Bial Biotech Inc. None of these companies were involved in any parts of preparing, drafting, and publishing this study.

### Funding Statement

This work was supported by a Foundation Scheme award to AD from the Canadian Institutes of Health Research, the Funai Foundation for Information Technology fellowship to MS, and by a Fonds de recherche du Quebec - Sante (FRQS) foreign post-doctoral training award to FM. UV has been funded by Estonian Research
Council's personal research funding start-up grant PSG759. This work was also financially supported by Parkinson Canada, the Canadian Consortium on
Neurodegeneration in Aging (CCNA), and the Canada First Research Excellence Fund (CFREF), awarded to McGill University for the Healthy Brains
for Healthy Lives (HBHL) program.

### Author Declarations

The study used ONLY openly available human data that were collected as part of the Adolescent Brain Cognitive Development study (https://abcdstudy.org/)

## References

1. Park B, Byeon K, Lee MJ, Chung C, Kim S, Morys F, et al. Whole-brain functional connectivity correlates of obesity phenotypes. Hum Brain Mapp. 2020;41:4912–4924.

2. Horstmann A, Busse FP, Mathar D, Muller K, Lepsien J, Schlogl H, et al. Obesity-Related Differences between Women and Men in Brain Structure and Goal-Directed Behavior. Front Hum Neurosci. 2011;5:58.

3. Morys F, García-García I, Dagher A. Is obesity related to enhanced neural reactivity to visual food cues? A review and meta-analysis. Soc Cogn Affect Neurosci. 2020. 2020. https://doi.org/10.1093/scan/nsaa113.

4. García-García I, Morys F, Dagher A. Nucleus accumbens volume is related to obesity measures in an age-dependent fashion. BioRxiv. 2019:773119.

5. García-García I, Michaud A, Dadar M, Zeighami Y, Neseliler S, Collins DL, et al. Neuroanatomical differences in obesity: meta-analytic findings and their validation in an independent dataset. Int J Obes. 2018:1.

6. Vainik U, Baker TE, Dadar M, Zeighami Y, Michaud A, Zhang Y, et al. Neurobehavioral correlates of obesity are largely heritable. Proc Natl Acad Sci. 2018;115:9312–9317.

7. Veit R, Kullmann S, Heni M, Machann J, Häring HU, Fritsche A, et al. Reduced cortical thickness associated with visceral fat and BMI. NeuroImage Clin. 2014;6:307–311.

8. Zhang B, Tian X, Tian D, Wang J, Wang Q, Yu C, et al. Altered regional gray matter volume in obese men: A structural MRI study. Front Psychol. 2017;8.

9. Gustafson D, Lissner L, Bengtsson C, Björkelund C, Skoog I. A 24-year follow-up of body mass index and cerebral atrophy. Neurology. 2004;63:1876–1881.

10. Raji CA, Ho AJ, Parikshak NN, Becker JT, Lopez OL, Kuller LH, et al. Brain structure and obesity. Hum Brain Mapp. 2010;31:353–364.

11. Rapuano KM, Laurent JS, Hagler DJ, Hatton SN, Thompson WK, Jernigan TL, et al. Nucleus accumbens cytoarchitecture predicts weight gain in children. Proc Natl Acad Sci U S A. 2020;117:26977–26984.

12. Rapuano KM, Zieselman AL, Kelley WM, Sargent JD, Heatherton TF, Gilbert-Diamond D. Genetic risk for obesity predicts nucleus accumbens size and responsivity to real-world food cues. 2017;114:160–165.

13. Opel N, Redlich R, Kaehler C, Grotegerd D, Dohm K, Heindel W, et al. Prefrontal gray matter volume mediates genetic risks for obesity. Mol Psychiatry. 2017;22:703–710.

14. Vainik U, García-García I, Dagher A. Uncontrolled eating: a unifying heritable trait linked with obesity, overeating, personality and the brain. Eur J Neurosci. 2019;50.

15. Locke AE, Kahali B, Berndt SI, Justice AE, Pers TH, Day FR, et al. Genetic studies of body mass index yield new insights for obesity biology. Nature. 2015;518:197–206.

16. Elks CE, Hoed M den, Zhao JH, Sharp SJ, Wareham NJ, Loos RJF, et al. Variability in the Heritability of Body Mass Index: A Systematic Review and Meta-Regression. Front Endocrinol (Lausanne). 2012;3.

17. Silventoinen K, Jelenkovic A, Sund R, Hur YM, Yokoyama Y, Honda C, et al. Genetic and environmental effects on body mass index from infancy to the onset of adulthood: an individual-based pooled analysis of 45 twin cohorts participating in the COllaborative project of Development of Anthropometrical measures in Twins (CODATwins) study. Am J Clin Nutr. 2016;104:371.

18. Yengo L, Sidorenko J, Kemper KE, Zheng Z, Wood AR, Weedon MN, et al. Meta-analysis of genome-wide association studies for height and body mass index in ∼ 700000 individuals of European ancestry. Hum Mol Genet. 2018;27:3641.

19. Herrera BM, Lindgren CM. The Genetics of Obesity. Curr Diab Rep. 2010;10:498.

20. Loos RJF, Yeo GSH. The genetics of obesity: from discovery to biology. Nat Rev Genet 2021 232. 2021;23:120–133.

21. Becker J, Burik CAP, Goldman G, Wang N, Jayashankar H, Bennett M, et al. Resource profile and user guide of the Polygenic Index Repository. Nat Hum Behav 2021 512. 2021;5:1744–1758.

22. Vainik U, Baker TE, Dadar M, Zeighami Y, Michaud A, Zhang Y, et al. Neurobehavioral correlates of obesity are largely heritable. Proc Natl Acad Sci. 2018:201718206.

23. Llewellyn CH, Van Jaarsveld CHM, Johnson L, Carnell S, Wardle J. Nature and nurture in infant appetite: analysis of the Gemini twin birth cohort. Am J Clin Nutr. 2010;91:1172–1179.

24. Garcia-Garcia I, Neseliler S, Morys F, Dadar M, Yau YHC, Scala SG, et al. Relationship between impulsivity, uncontrolled eating and body mass index: a hierarchical model. Int J Obes (Lond). 2022;46:129–136.

25. Morys F, Dadar M, Dagher A. Association between mid-life obesity, its metabolic consequences, cerebrovascular disease and cognitive decline. J Clin Endocrinol Metab. 2021. 2 March 2021. https://doi.org/10.1210/clinem/dgab135.

26. Thaler JP, Yi CX, Schur EA, Guyenet SJ, Hwang BH, Dietrich MO, et al. Obesity is associated with hypothalamic injury in rodents and humans. J Clin Invest. 2012;122:153.

27. Douglass JD, Dorfman MD, Thaler JP. Glia: silent partners in energy homeostasis and obesity pathogenesis. Diabetologia. 2017;60:226–236.

28. Tomassoni D, Martinelli I, Moruzzi M, Di Bonaventura MVM, Cifani C, Amenta F, et al. Obesity and Age-Related Changes in the Brain of the Zucker Lepr fa/fa Rats. Nutrients. 2020;12.

29. Namavar MR, Raminfard S, Jahromi ZV, Azari H. Effects of high-fat diet on the numerical density and number of neuronal cells and the volume of the mouse hypothalamus: a stereological study. Anat Cell Biol. 2012;45:178.

30. Bocarsly ME, Fasolino M, Kane GA, Lamarca EA, Kirschen GW, Karatsoreos IN, et al. Obesity diminishes synaptic markers, alters microglial morphology, and impairs cognitive function. Proc Natl Acad Sci. 2015;112:15731–15736.

31. Oginsky MF, Maust JD, Corthell JT, Ferrario CR. Enhanced cocaine-induced locomotor sensitization and intrinsic excitability of NAc medium spiny neurons in adult but not in adolescent rats susceptible to diet-induced obesity. Psychopharmacology (Berl). 2016;233:773–784.

32. Bouret SG, Gorski JN, Patterson CM, Chen S, Levin BE, Simerly RB. Hypothalamic Neural Projections Are Permanently Disrupted in Diet-Induced Obese Rats. Cell Metab. 2008;7:179–185.

33. Levin BE, Dunn-Meynell AA, Banks WA. Obesity-prone rats have normal blood-brain barrier transport but defective central leptin signaling before obesity onset. Am J Physiol - Regul Integr Comp Physiol. 2004;286:143–150.

34. Richardson TG, Sanderson E, Elsworth B, Tilling K, Smith GD. Use of genetic variation to separate the effects of early and later life adiposity on disease risk: mendelian randomisation study. BMJ. 2020;369.

35. Casey BJ, Cannonier T, Conley MI, Cohen AO, Barch DM, Heitzeg MM, et al. The Adolescent Brain Cognitive Development (ABCD) study: Imaging acquisition across 21 sites. Dev Cogn Neurosci. 2018;32:43–54.

36. Garavan H, Bartsch H, Conway K, Decastro A, Goldstein RZ, Heeringa S, et al. Recruiting the ABCD sample: Design considerations and procedures. Dev Cogn Neurosci. 2018;32:16–22.

37. Barch DM, Albaugh MD, Avenevoli S, Chang L, Clark DB, Glantz MD, et al. Demographic, physical and mental health assessments in the adolescent brain and cognitive development study: Rationale and description. Dev Cogn Neurosci. 2018;32:55–66.

38. Growth Charts - Clinical Growth Charts. https://www.cdc.gov/growthcharts/clinical_charts.htm. Accessed 9 August 2021.

39. Hagler DJ, Hatton SN, Cornejo MD, Makowski C, Fair DA, Dick AS, et al. Image processing and analysis methods for the Adolescent Brain Cognitive Development Study. Neuroimage. 2019;202:116091.

40. Desikan RS, Ségonne F, Fischl B, Quinn BT, Dickerson BC, Blacker D, et al. An automated labeling system for subdividing the human cerebral cortex on MRI scans into gyral based regions of interest. Neuroimage. 2006;31:968–980.

41. Fischl B. FreeSurfer. Neuroimage. 2012;62:774–781.

42. Hagler DJ, Ahmadi ME, Kuperman J, Holland D, McDonald CR, Halgren E, et al. Automated white-matter tractography using a probabilistic diffusion tensor atlas: Application to temporal lobe epilepsy. Hum Brain Mapp. 2009;30:1535– 1547.

43. Fortin J-P, Cullen N, Sheline YI, Taylor WD, Aselcioglu I, Cook PA, et al. Harmonization of cortical thickness measurements across scanners and sites. Neuroimage. 2018;167:104–120.

44. Choi S, O’Reilly P. PRSice-2: Polygenic Risk Score software for biobank-scale data. Gigascience. 2019;8.

45. Jernigan TL, Brown TT, Hagler DJ, Akshoomoff N, Bartsch H, Newman E, et al. The Pediatric Imaging, Neurocognition, and Genetics (PING) Data Repository. Neuroimage. 2016;124:1149.

46. Vainik U, Dagher A, Dubé L, Fellows LK. Neurobehavioural correlates of body mass index and eating behaviours in adults: A systematic review. Neurosci Biobehav Rev. 2013;37:279–299.

47. Robinson E, Roberts C, Vainik U, Jones A. The psychology of obesity: An umbrella review and evidence-based map of the psychological correlates of heavier body weight. Neurosci Biobehav Rev. 2020;119:468–480.

48. Whiteside SP, Lynam DR, Miller JD, Reynolds SK. Validation of the UPPS impulsive behaviour scale: A four-factor model of impulsivity. Eur J Pers. 2005;19:559–574.

49. Watts AL, Smith GT, Barch DM, Sher KJ. Factor structure, measurement and structural invariance, and external validity of an abbreviated youth version of the UPPS-P Impulsive Behavior Scale. Psychol Assess. 2019;32:336.

50. Booth C, Spronk D, Grol M, Fox E. Uncontrolled eating in adolescents: The role of impulsivity and automatic approach bias for food. Appetite. 2018;120:636–643.

51. VanderBroek-Stice L, Stojek MK, Beach SRH, vanDellen MR, MacKillop J. Multidimensional assessment of impulsivity in relation to obesity and food addiction. Appetite. 2017;112:59–68.

52. Ronan L, Alexander-Bloch A, Fletcher PC. Childhood Obesity, Cortical Structure, and Executive Function in Healthy Children. Cereb Cortex. 2019;30:2519–2528.

53. Luciana M, Bjork JM, Nagel BJ, Barch DM, Gonzalez R, Nixon SJ, et al. Adolescent neurocognitive development and impacts of substance use: Overview of the adolescent brain cognitive development (ABCD) baseline neurocognition battery. Dev Cogn Neurosci. 2018;32:67–79.

54. Akshoomoff N, Beaumont JL, Bauer PJ, Dikmen SS, Gershon RC, Mungas D, et al. VIII. NIH Toolbox Cognition Battery (CB): composite scores of crystallized, fluid, and overall cognition. Monogr Soc Res Child Dev. 2013;78:119–132.

55. Akshoomoff N, Newman E, Thompson WK, McCabe C, Bloss CS, Chang L, et al. The NIH Toolbox Cognition Battery: results from a large normative developmental sample (PING). Neuropsychology. 2014;28:1–10.

56. Akshoomoff N, Brown TT, Bakeman R, Hagler DJ. Developmental differentiation of executive functions on the NIH Toolbox Cognition Battery. Neuropsychology. 2018;32:777–783.

57. R Core Team. R: The R Project for Statistical Computing. 2013.

58. Benjamini Y, Hochberg Y. Controlling the False Discovery Rate: A Practical and Powerful Approach to Multiple Testing. J R Stat Soc Ser B. 1995;57:289– 300.

59. Schäfer T, Ecker C. fsbrain: an R package for the visualization of structural neuroimaging data. BioRxiv. 2020:2020.09.18.302935.

60. Mesulam MM. From sensation to cognition. Brain. 1998;121:1013–1052.

61. Vainik U, Paquola C, Wang X, Zheng Y, Bernhardt B, Misic B, et al. Heritability of cortical morphology reflects a sensory-fugal plasticity gradient. BioRxiv. 2020:2020.11.03.366419.

62. Smith SM, Vidaurre D, Alfaro-Almagro F, Nichols TE, Miller KL. Estimation of brain age delta from brain imaging. Neuroimage. 2019;200:528–539.

63. Zeighami Y, Dadar M, Daoust J, Pelletier M, Biertho L, Bouvet-Bouchard L, et al. Impact of Weight Loss on Brain Age: Improved Brain Health Following Bariatric Surgery. 2021. 12 December 2021.

64. Rosseel Y. lavaan: An R Package for Structural Equation Modelinge human forearm during rythmic exercise. J Stat Softw. 2012;48:1–36.

65. Llewellyn C, Wardle J. Behavioral susceptibility to obesity: Gene-environment interplay in the development of weight. Physiol Behav. 2015;152:494–501.

66. Dohle S, Diel K, Hofmann W. Executive functions and the self-regulation of eating behavior: A review. Appetite. 2018;124:4–9.

67. García-García I, Michaud A, María ·, Jurado Á, Dagher A, Morys F. Mechanisms linking obesity and its metabolic comorbidities with cerebral grey and white matter changes. Rev Endocr Metab Disord 2021. 2022;1:1–11.

68. Corbetta M, Shulman GL. Control of goal-directed and stimulus-driven attention in the brain. Nat Rev Neurosci. 2002;3:201–215.

69. Dagher A. Functional brain imaging of appetite. Trends Endocrinol Metab. 2012;23:250–260.

70. Wang Q, Chen C, Cai Y, Li S, Zhao X, Zheng L, et al. Dissociated neural substrates underlying impulsive choice and impulsive action. Neuroimage. 2016;134:540–549.

71. Sydnor VJ, Larsen B, Bassett DS, Alexander-Bloch A, Fair DA, Liston C, et al. Neurodevelopment of the association cortices: Patterns, mechanisms, and implications for psychopathology. Neuron. 2021;109:2820–2846.

72. Park B yong, Park H, Morys F, Kim M, Byeon K, Lee H, et al. Inter-individual body mass variations relate to fractionated functional brain hierarchies. Commun Biol 2021 41. 2021;4:1–12.

73. Calvo D, Galioto R, Gunstad J, Spitznagel MB. Uncontrolled eating is associated with reduced executive functioning. Clin Obes. 2014;4:172–179.

74. Prunell-Castañé A, Jurado MÁ, García-García I. Clinical binge eating, but not uncontrolled eating, is associated with differences in executive functions: Evidence from meta-analytic findings. Addict Behav Reports. 2021;13:100337.

75. McClelland J, Dalton B, Kekic M, Bartholdy S, Campbell IC, Schmidt U. A systematic review of temporal discounting in eating disorders and obesity: Behavioural and neuroimaging findings. Neurosci Biobehav Rev. 2016;71:506–528.

76. Mobbs O, Crépin C, Thiéry C, Golay A, Van der Linden M. Obesity and the four facets of impulsivity. Patient Educ Couns. 2010;79:372–377.

77. Sharkey RJ, Bourque J, Larcher K, Mišić B, Zhang Y, Altınkaya A, et al. Mesolimbic connectivity signatures of impulsivity and BMI in early adolescence. Appetite. 2019;132:25–36.

78. Nederkoorn C, Braet C, Van Eijs Y, Tanghe A, Jansen A. Why obese children cannot resist food: the role of impulsivity. Eat Behav. 2006;7:315–322.

79. Llewellyn CH, Van Jaarsveld CHM, Boniface D, Carnell S, Wardle J. Eating rate is a heritable phenotype related to weight in children. Am J Clin Nutr. 2008;88:1560–1566.

80. Fildes A, Van Jaarsveld CHM, Llewellyn CH, Fisher A, Cooke L, Wardle J. Nature and nurture in children’s food preferences. Am J Clin Nutr. 2014;99:911–917.

81. Herle M, Smith AD, Kininmonth A, Llewellyn C. The Role of Eating Behaviours in Genetic Susceptibility to Obesity. Curr Obes Rep. 2020;9:512–521.

82. Llewellyn CH, Fildes A. Behavioural Susceptibility Theory: Professor Jane Wardle and the Role of Appetite in Genetic Risk of Obesity. Curr Obes Rep. 2017;6:38–45.

83. Nederkoorn C, Dassen FCM, Franken L, Resch C, Houben K. Impulsivity and overeating in children in the absence and presence of hunger. Appetite. 2015;93:57–61.

